# Temporal trends in *Plasmodium vivax* diversity in eastern Cambodia evidence declining transmission

**DOI:** 10.64898/2026.03.03.26346840

**Authors:** Rotha Eam, Kian Soon Hoon, Edwin Sutanto, Anjana Rai, Hidayat Trimarsanto, Angela Rumaseb, Sopheany Thin, Sreyneat Hor, Chhea Chhorvann, Tol Bunkea, Ric N Price, Jean Popovici, Sarah Auburn

## Abstract

**Background:** Elimination of *Plasmodium vivax* is challenging due to its dormant liver stages (hypnozoites), which can reactivate weeks or months after the primary infection, causing relapses and ongoing transmission of the parasite. Despite these challenges, *P. vivax* clinical case numbers have declined over the past decade in Cambodia. We used parasite genotyping to assess whether the decline in case numbers was reflected in parasite diversity and relatedness as a proxy to transmission.

**Methods:** Genotyping was conducted on 182 symptomatic *P. vivax* isolates collected in eastern Cambodia in 2014, 2015, 2019 and 2023. A panel of 93 microhaplotype markers (vivaxGEN panel) was genotyped using Illumina sequencing. Population genetic measures were applied to determine infection diversity and relatedness (identity-by-descent (IBD)) each year.

**Results:** The genetic results correlated well with clinical case numbers for the study years. The percentage of polyclonal infections was 5% in 2023 compared to 22-48% in earlier years (p<0.05) suggesting substantial reduction in superinfection and aligning with accelerated primaquine use in 2021. The cases in 2023 also had the highest percentage of infections with IBD >0.95 with one or more other infections (81.4% versus 8.9-10.8% in 2014-2019) indicative of inbreeding following population bottlenecking. In 2019, there was a spike in polyclonal infections (48%) and population diversity following local interruption of critical malaria control services.

**Conclusions:** Our findings illustrate the potential of microhaplotype genotyping to inform on *P. vivax* transmission to assess intervention efficacy. In eastern Cambodia, the data provides evidence to support of widespread use of radical cure for patients with *P. vivax* malaria.

## Background

*Plasmodium vivax* is the leading cause of malaria in the Asia-Pacific region, where over 80% of the global burden occurs (1)(2). *P. vivax* elimination is challenging due to the parasite’s ability to form undetectable dormant liver stages (hypnozoites), that can reactivate weeks or months after the primary infection, causing recurrent episodes of malaria known as relapses (3). In populations with high transmission rates, up to 85% of recurrences are caused by relapses, contributing substantially to the incidence of infection and the ongoing transmission of the parasite (4). The WHO recommends the radical cure of patients with *P. vivax* malaria using a combination of schizonticidal (chloroquine or artemisinin-based based combination therapies) plus a hypnozoitocidal agent, such as primaquine and tafenoquine, to kill the dormant liver stages (1). Primaquine treatment is usually administered over 7-14 days but this prolonged course is associated with poor adherence and effectiveness (1)(5)(6). Furthermore, patients with host genetic variants in the gene encoding Cytochrome P450 2D6 (CYP2D6) enzyme may not metabolize primaquine or tafenoquine effectively and experience treatment failure as a result (7). National malaria control and elimination programs need to maintain diligent surveillance of the effectiveness of radical cure and other interventions to ensure that they stay on track to meet their malaria elimination goals (8). However, monitoring transmission reduction can be challenging owing to hidden reservoirs in the liver as well as a newly discovered hidden splenic reservoir (9). Asymptomatic and subpatent infections further complicate efforts to monitor the burden of infection.

Malaria genomic epidemiology has potential to provide insights on transmission intensity that complement traditional measures of incidence (10)(11). Genetic data can be leveraged to characterize within-host *P. vivax* diversity and thereby clarify the processes of shaping parasite transmission. The presence of multiple genetically distinct clones within a single infection often reflects either superinfection from repeated infectious bites or co-transmission of mixed parasite populations by a single mosquito, providing estimates of the force of infection (11). At the population level, identity-by-descent (IBD) patterns further complement these measures by capturing recent ancestry among parasites (12). Elevated IBD values are characteristic of populations that have experienced reduced recombination opportunities due to declining transmission and progressive bottlenecking of the parasite reservoir. These combined signatures enable characterization of the epidemiological dynamics of *P. vivax* in settings progressing toward elimination. A range of *P. vivax* genotyping methods are available including capillary sequencing of microsatellite markers or next-generation sequencing (NGS) of SNPs or microhaplotypes (amplicons containing multiple, highly informative SNPs) (13) (14). NGS-based amplicon sequencing is the most common method for malaria molecular surveillance since it provides high throughput and cost-effective genotyping of large marker panels that can inform on multiple epidemiological use cases including monitoring transmission (15). To date, malaria molecular surveillance in the Asia-Pacific has largely focused on *P. falciparum*, but the increasing predominance of *P. vivax* cases and recent availability of a *P. vivax* microhaplotype panel has refocused surveillance efforts on this species (16). The microhaplotype panel is globally applicable with high diversity and accuracy to quantify IBD in *P. vivax* infections from the Asia-Pacific (16). The panel therefore has potential to capture spatial and temporal trends in *P. vivax* transmission.

All countries in the Greater Mekong Subregion, including Cambodia, have set ambitious targets to eliminate all malaria by 2030, regardless of the *Plasmodium* species, but several endemic countries are facing the challenge of rising proportion of malaria due to *P. vivax* (17) (18). In 2019, the Cambodian National Malaria Elimination Program introduced the radical cure of *P vivax* with primaquine after glucose-6-phosphate dehydrogenase (G6PD) to accelerate the control and elimination of *P. vivax*. In 2021, after a period of pilot testing in selected health centers, primaquine was rolled out across all endemic areas of the country (8). Since then, the number of reported malaria cases has steadily declined, including those caused by *P. vivax*.

In this study we investigate the use of microhaplotype genotyping to capture diversity, relatedness and associated transmission of *P. vivax* isolates collected in eastern Cambodia, to confirm substantial changes in *P. vivax* epidemiology over a 10-year period.

## Methods

### Study sites and sample collection

A total of 182 venous blood samples were collected from patients with uncomplicated *P. vivax* malaria presenting to health clinics at two provinces in eastern Cambodia: Rattanakiri (Banlung district) and Mondulkiri (Keo Seima, Figure 1a). Symptomatic patients seeking treatment were recruited passively in 2014, 2015, 2019 and 2023 as summarized in Supplementary Table 1. All patients confirmed to have *P. vivax* parasites by rapid diagnostic testing, performed by Village Malaria Workers or Health Center staff, were offered to participate. Upon consent, a venous blood sample was collected, and patients were treated by the health workers according to the national treatment guidelines. Rattanakiri and Mondulkiri provinces have similar trends in reported *P. vivax* case counts between 2014 and 2023, both demonstrating a spike in cases in 2019, followed by a drop in cases in 2023 (Figure 1b).

**Figure 1.**
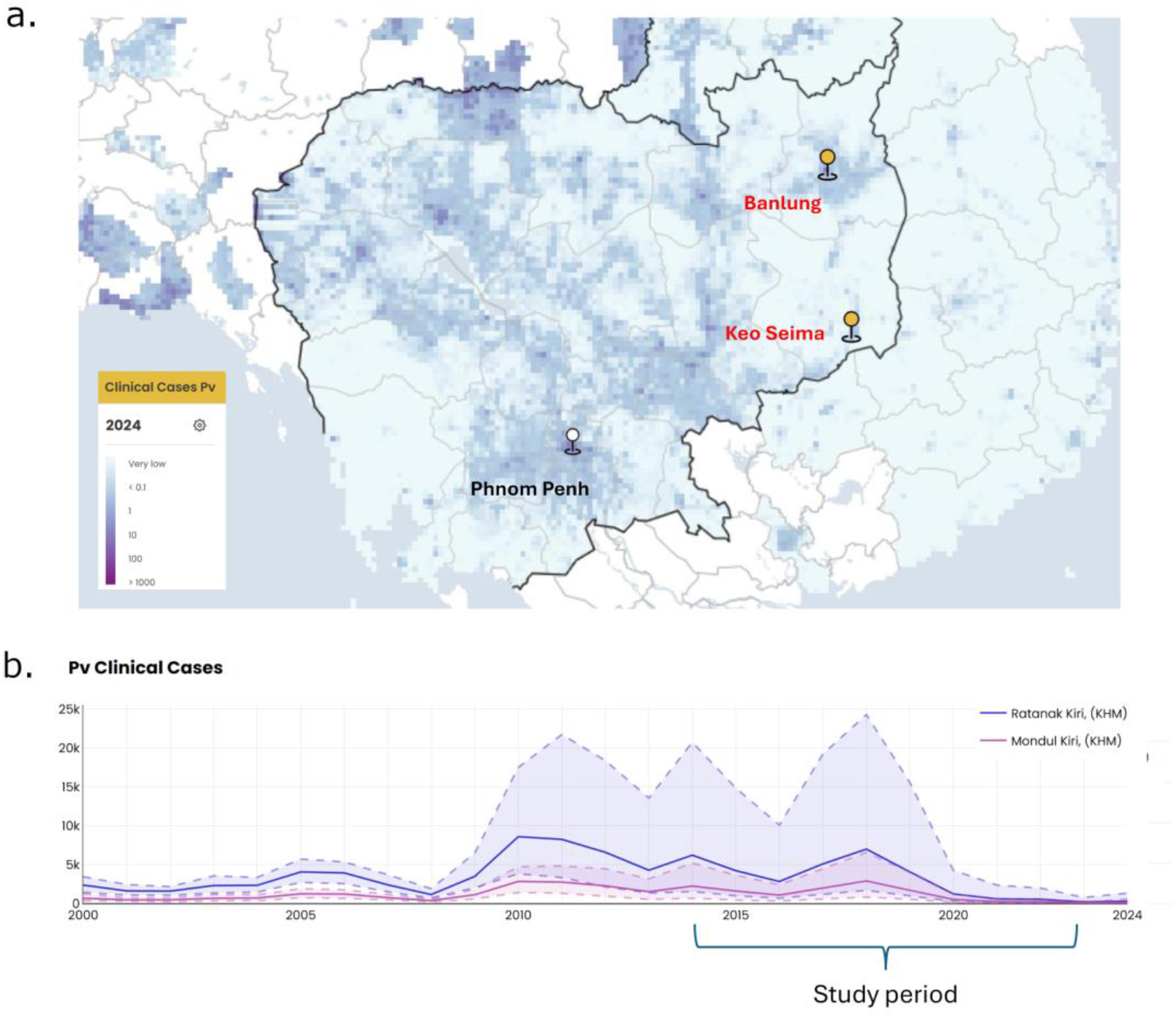
Study site locations and *P. vivax* clinical case counts. Panel a) indicates the location of the study sites, Banlung and Keo Seima, where *P. vivax* infected patients were enrolled relative to the capital city (Phnom Penh). The study clinics are shown on an incidence density map generated by the Malaria Atlas Project (19). Panel b) presents the number of newly diagnosed *P. vivax* cases per 1,000 population for Rattanakiri and Mondulkiri Provinces between 2000 and 2024 with demarcation of the study period (2014-2023). The dashed lines and shaded areas depict the 95% confidence interval for the given year. The clinical case numbers in panel b) are also derived from the Malaria Atlas Project (19).

### DNA extraction and species confirmation

DNA extraction was performed using the QIAamp DNA Mini Kit and *P. vivax* monoinfection was confirmed by species-specific real-time PCR targeting *cytB* enabling the identification of P. *falciparum, P. vivax, P. ovale* and *P. malariae* (20).

### rhAmpSeq library preparation and sequencing

*P. vivax* genotyping was conducted using the vivaxGEN microhaplotype panel which comprises 98 markers including 93 microhaplotypes (16). In brief, library preparation was performed using a rhAmpSeq (Integrated DNA Technologies) workflow, comprising a first round of multiplex PCR to amplify the 98 markers in each sample, and a secondary PCR to incorporate the indexes. The final library was purified using beads and checked for quality based on fragment size and qPCR using Quant Studio real-time PCR. Illumina sequencing was conducted using a MiniSeq Mid Output Kit (300 cycles; Illumina, USA), generating 150 bp paired end reads on a MiniSeq machine. Up to 96 samples were multiplexed in each sequencing run.

### Variant calling and population genetic analysis

Microhaplotype variant calling was conducted using the vivaxGEN-Microhaps pipeline (https://github.com/vivaxgen/MicroHaps). The pipeline uses the *cutadapt* software (21) to remove adapter and primer sequences and perform quality trimming at the right-hand end of the reads. Reads were then error-corrected, denoised, and subsequently merged with *DADA2* software to generate amplicon sequence variants (21)(22). Custom R script was used to post-process *DADA2* output, including filtering for valid genotype calls. Haplotypes with occurrence less than 2% or 5 read pairs per sample were filtered out, and a threshold of 75 (80%) genotype calls was required for successful genotyping of a sample. One of the microhaplotype markers (PvP01_04_v2:361776-361913:64721) failed and was excluded from subsequent analysis.

Sample expected heterozygosity was calculated by summing up the sum of squared allele frequencies per marker per sample divided by the number of markers per sample. Within-host infection diversity was determined using the effective multiplicity of infection (eMOI) metric using *MOIRE* software (version 3.5.0) with the following parameters: burnin=1000; sample_per_chain=1000; pt_chains=seq(0.5, 1, length.out = 40); r_alpha = 3; r_beta = 3 (23). *MOIRE* was also used to inform on the probability that each sample was polyclonal, using a threshold of >50% probability to classify infections as polyclonal. The R-based *paneljudge* package was used to measure the heterozygosity/diversity and effective cardinality of the marker set, with restriction to monoclonal samples (24). The relatedness, as measured by identity-by-descent (IBD) between-host infections was performed using *Dcifer* (version 1.3.0) (25). Network plots illustrating the IBD between infections were generated using the R-based *ggnetwork* package (26). Samples were assigned to one of three categorical IBD groupings based on the maximum IBD observed with the other infections within the given year group as follows: low-moderate = IBD 0-0.25, high = IBD 0.25-0.95, and identical/near-identical = IBD >0.95. Identity-by-state (IBS) was measured using the R-based *ape* package (version 5.8.1) to calculate genetic distance and plotted as neighbour-joining trees and principal coordinate plots (PCoA) (27). All plots were prepared using the *ggplot2* package (28). F_ST_ (Weir & Cockerham’s θ) (29) and standardized F_ST_ (Meirmans & Hedrick’s G’’_ST_ (30) between Rattanakiri and Mondulkiri provinces were calculated using hierfstat (31) and mmod (32), respectively.

### Statistical tests

R software (version 4.5.2) was used to conduct statistical tests (33). Annual variation in the proportion of polyclonal infections were determined using N-1 Chi-squared test (34). The proportion of infections within each IBD group was determined using Chi-squared test. Multiple comparisons were accounted for using Benjamini-Hochberg correction. Differences in the distributions of eMOI and expected heterozygosity by year were assessed using a Kruskal-Wallis test (35). If the Kruskal-Wallis test was significant, additional post-hoc analysis was done using Conover-Iman test (36), with false discovery rate controlled using Benjamini-Hochberg method (37). P-values below 0.05 were considered significant.

### Ethical approval

Genetic analysis of *P. vivax* was approved by the National Ethics Committee for Health Research of the Ministry of Health of Cambodia (#364-NECHR, #038-NECHR and #197-NECHR).

### Code availability

The multiallelic microhaplotype calls were generated using the vivaxGEN pipeline available on GitHub via the link https://github.com/vivaxgen/MicroHaps.

## Results

### High quality data generated across 2014-2023

A total of 182 samples were collected from patients with *P. vivax* malaria between 2014 and 2023 (Supplementary Data 1). Majority of patients were males (75%, 137/182) and aged more than 16 years old (65%, 118/182) (Supplementary Table 1). Overall, 96% (175/182) of isolates were successfully genotyped and included in the population genetic analyses, with 37-50 isolates available for each year. One of the 93 microhaplotype markers (marker 64721, PvP01_04_v2:361776-361913:64721) failed in all 175 high-quality samples and was excluded from the analyses. The majority of the remaining 92 microhaplotype markers displayed moderate to high marker diversity (median of 0.696 in 2014, 0.712 in 2015, 0.706 in 2019, 0.672 in 2023) and effective cardinalities (median of 3.29 in 2014, 3.47 in 2015, 3.40 in 2019, 3.05 in 2023), confirming potential to discriminate distinct clones and capture IBD (Supplementary Figure 1, Supplementary Data 2).

### Significant changes in within-host diversity in 2019 and 2023

Within-host diversity was first assessed using polyclonal infection frequencies. The prevalence of polyclonal infections was high in 2014 (41%,15/37), 2015 (22%, 10/45) and 2019 (48%, 24/50) but fell significantly to 5% (2/43) in 2023 (p < 0.001) (Figure 2a, Supplementary Table 2 and Data 3). The relatedness of within-host infections (expressed as eMOI) was also used to determine the complexity of parasite infections within the host. Significant differences in eMOI were oberved over the study period (p = 2.042 × 10^-5^) (Figure 2b). Post-hoc analysis revealed notably an increase of eMOI between 2015 and 2019 (p = 0.011) followed by a sharp reduction in 2023 (p = 2.507 × 10^-6^).

**Figure 2:**
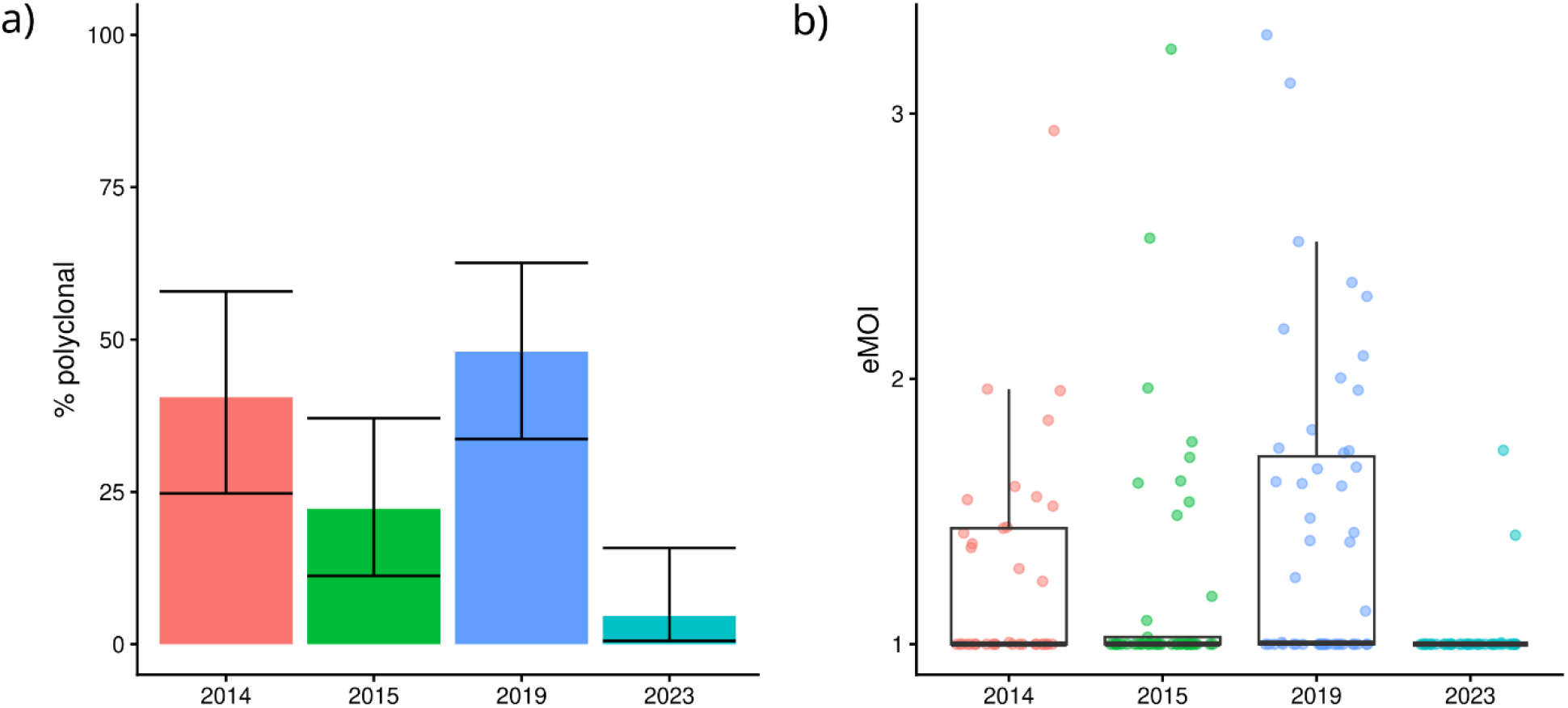
Within-host diversity trends over time. Panel a) presents the prevalence of polyclonal infections for each year, with error bars representing the 95% confidence intervals. Panel b) presents the effective multiplicity of infection (eMOI) for each year, with dots representing the individual eMOI.

### Increasing relatedness between host infections over time

Analysis of the relatedness between infections within each year’s group revealed a trend of increasing IBD across the years (Figure 3, Supplementary Data 4). A large proportion of the infections collected in 2023 exhibited relatedness close to 1. On applying categorical groupings to determine the maximum IBD observed for each infection as low (<25%), moderate (25-95%) or high (>95%), significant differences were observed in the proportion of identical/near-identical infections between the years (χ^2^ = 274.17, df = 6, p-value < 2.2 × 10^-16^). Using consecutive year comparisons of the proportions of samples in each IBD category, there was no significant difference between 2014 and 2015 (χ^2^ = 3.568, df = 2, p = 0.168), but significant differences were observed between 2015 and 2019 (χ^2^ = 15.902, df = 2, p = 5.284 × 10^-4^), and between 2019 and 2023 (χ^2^ = 110.77, df = 2, p < 2.2 × 10^-16^), reflecting increasing proportions of identical infections over time.

**Figure 3:**
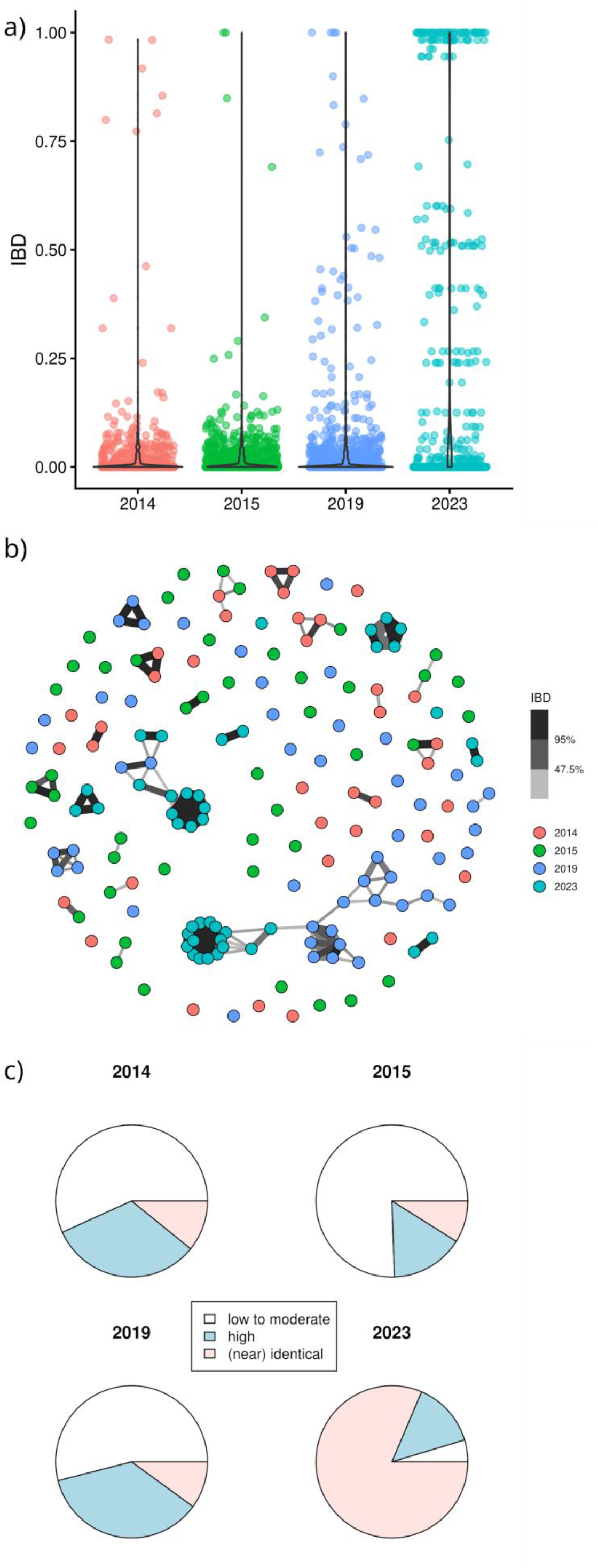
IBD-based relatedness between host infections. Panel a) presents the distribution of IBD values between infections in the given years. Panel b) presents a network plot illustrating sample connections across IBD thresholds ranging from 25% to 95%. Each circle represents an infection, with colour-coding by year of collection. The lines connecting infections reflect IBD, whereby the darkness and thickness of the lines reflect the IBD level (darker and thicker lines represent higher IBD level). Panel c) presents pie charts illustrating the maximum IBD observed for each infection with categorical groupings of low-moderate (IBD<25%), high (25%-95%) and identical/near-identical (>95%).

Measures of identity-by-state (IBS) between the infections using a genetic distance matrix applied to the monoclonal sample set (n=124) confirmed the temporal trends observed with IBD analysis (Supplementary Figure 2). The IBS-based neighbor-joining plots also demonstrated low differentiation between the cases from Mondulkiri and Rattanakiri, justifying pooling of the cases across the two provinces. Among monoclonal infections and excluding multiply represented clones (which can potentially bias measures), low F_ST_ (0.018) and standardized F_ST_ (0.086) values were observed between Mondulkiri (n=53) and Rattanakiri (n=55), providing additional confirmation of low differentiation between the provinces. When multiply represented clones were included, the F_ST_ (0.028) and standardized F_ST_ values (0.109) between Mondulkiri (n=67) and Rattanakiri (n=57) remained low.

### Low population-level genetic diversity in 2023

Population-level genetic diversity was examined using measures of expected heterozygosity applied to all isolates. The expected heterozygosity determines the probability that any two infections taken from the given population will have different multilocus genotypes and ranges from 0 (0% probability of difference) to 1 (100% probability of difference). There was significant variation in the population diversity over the study period, the mean heterozygosity ranging from 0.009 to 0.074 (p = 4.292 × 10^-7^) (Figure 4, Supplementary Data 3) with samples from 2023 showing the lowest heterozygosity.

**Figure 4.**
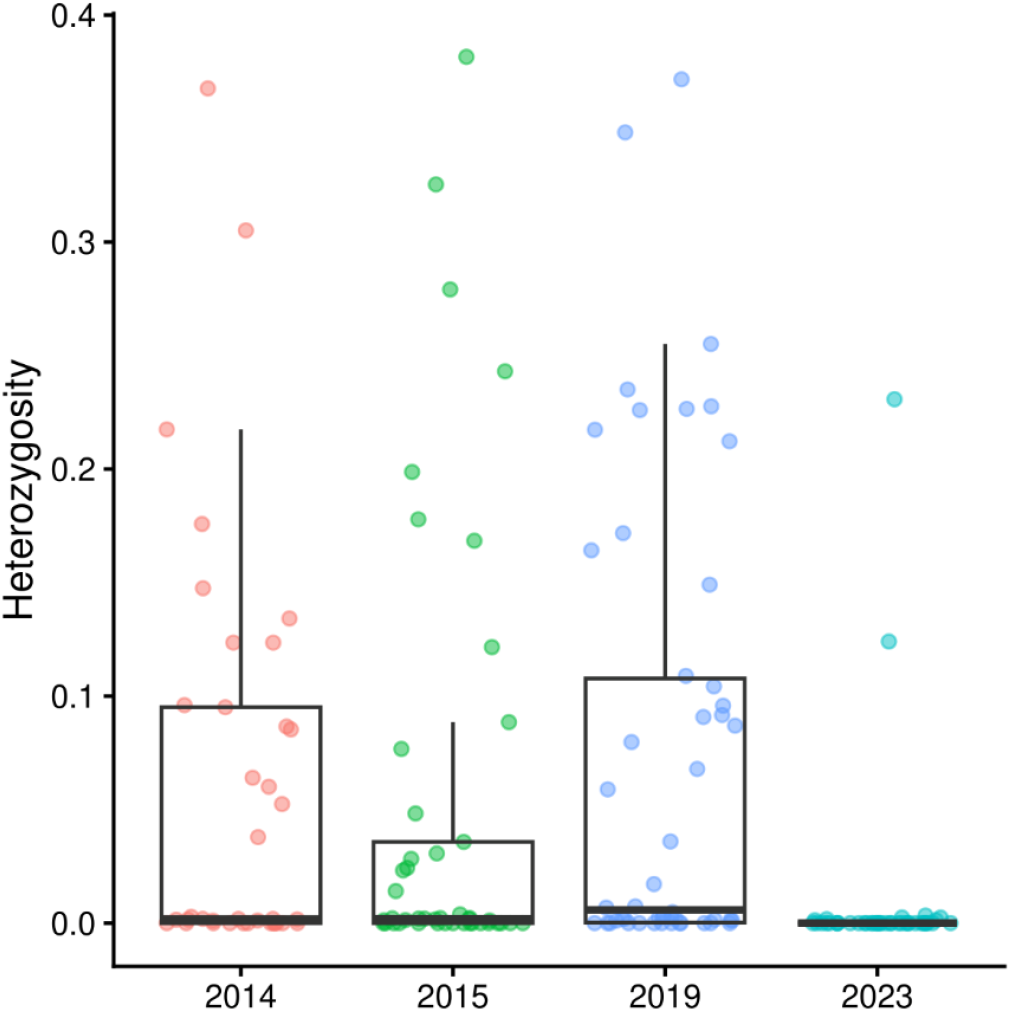
Temporal trends in genetic diversity. Boxplot distributions of the average expected heterozygosity for each sample within the given year group.

## Discussion

Our temporal study of *P. vivax* genetic epidemiology in eastern Cambodia between 2014 and 2023 reveals fluctuations in diversity that are highly consistent with the incidence of clinical cases over the same period. The trends in genetic diversity also align with major changes in the local malaria intervention strategies including a marked reduction in diversity and increased inbreeding following the implementation of primaquine radical cure, suggesting that introduction of intervention reduced transmission. These findings provide important proof of concept for the utility of genetic surveillance using symptomatic *P. vivax* cases to inform on the magnitude of the local reservoir of infection.

The potential for genetic epidemiology to inform parasite transmission is well described, but few studies have shown a direct application in temporal investigations over multiple years, particularly for *P. vivax* (38)(39). While the relationship between *P. falciparum* diversity and transmission is relatively direct, differences in parasite biology complicate the interpretation of data generated from *P. vivax* (40). For example, the impact of *P. vivax* hypnozoites in sustaining highly related infections is unclear. Previous temporal studies of *P. vivax* diversity have mostly used panels of up to 14 microsatellite markers, which lack the resolution to capture relatedness, as measured by IBD, accurately (41). Our study provides the first description of temporal trends in *P. vivax* relatedness using a sufficiently powered microhaplotype panel to measure IBD with low error. We demonstrate a strong correlation of within-host and population diversity and moderate correlation of genetic relatedness with clinical case counts of *P. vivax* over the study years in eastern Cambodia.

The temporal genetic trends in polyclonality, within-host and population diversity correlated with both the annual incidence of *P. vivax* reported during the study period and the major intervention changes undertaken in Cambodia. Relative to 2015 (22%), the prevalence of polyclonal infections showed a significant increase in 2019 (48%), followed by a significant decrease in 2023 (5%). Similar trends were observed in the within-host and population-level infection diversity. The trends suggest that superinfection, co-transmission and associated out-crossing levels had a marked increase in 2019 followed by a marked decrease in 2023. In alignment with these trends, the number of malaria cases in Mondulkiri and Rattanakiri declined in 2016, but rose substantially in 2017 and 2018, before dropping again in 2020 (42). The rising trends in 2017 and 2018 occurred after many Village Malaria Workers were dismissed and activities supported by The Global Fund were suspended between 2015 and 2016 (43)(44). In 2018 an intensification plan was initiated, and the subsequent number of malaria cases decreased from 3.9 per 1000 population at risk in 2018 to 1.9 per 1000 in 2019. Finally, the gradual introduction of primaquine in addition to a blood-stage schizonticide in 2019 and full rollout in the country in 2021 most likely had a great impact on reducing the reservoir of *P. vivax* parasites. Accordingly, a sharp decrease in cases and the lowest polyclonality rates and within-host diversity were observed in samples collected in 2023.

Polyclonality and within-host diversity were directly correlated with case counts, whereas relatedness (IBD) showed a less direct association. Despite a spike in reported cases in 2019, there was a consistent rise in relatedness over the study years, with increasingly large networks of identical or near-identical infections. A potential hypothesis for the limited impact of the rise in cases in 2017-18 on infection relatedness in 2019 is that a selection of lineages expanded rapidly and were potentially sustained by relapses from the liver stage reservoir. In this scenario, within-host and population diversity would be more sensitive than relatedness as proxies to distinguish expansion versus bottleneck dynamics.

The impact of an asymptomatic reservoir, in the liver, peripheral blood and spleen, on the observed diversity trends in our study is also unclear. Our study used *P. vivax* cases collected from symptomatic patients. A previous study in the meso-endemic setting of Papua Indonesia found no differences in *P. vivax* population-level diversity, but significant differences in polyclonality between passive (clinical) and actively (asymptomatic and sub-patent or symptomatic) detected cases (45). However, the Papuan study was based on 9 microsatellite markers and hence had low genetic resolution and did not investigate IBD. Further studies are needed to understand the differences between symptomatic and asymptomatic patients in low-endemic settings such as Cambodia. Additional research is also needed to identify optimal sample size and catchment areas for genetic surveillance to gauge transmission intensity. Clear guidelines are available for sampling for drug resistance detection, but other malaria molecular use cases are less clear (46). Lastly, it is unclear whether patients’ demographics, such as age and gender, may impact *P. vivax* diversity. This is a potential limitation of our study since there were significant differences in patient demographics between years that likely reflect changes in malaria epidemiology associated with declining transmission.

Another limitation of our study is the absence of metadata on patient home locality and occupation, which could have potentially identified any risk factors for malaria acquisition and supported mapping of transmission networks amongst the highly related strains observed in 2019 and 2023. As such, we cannot infer any hypothesis on spatial or behavioral factors that could have contributed to infection risk or shared strains between different patients. Future studies should be performed to better document any relations between parasite relatedness and malaria exposure.

In summary, our study provides proof of concept on the use of microhaplotype genotyping to inform on temporal trends in genetic diversity and associated transmission dynamics. However, further research is needed to explore optimal sampling methods in different endemic settings and extrapolation of our findings to the asymptomatic reservoir. Using modest sampling of clinical cases, we highlight evidence from genetic analyses that complement routine reporting of clinical case and supports the efficacy of primaquine implementation to reduce *P. vivax* transmission in eastern Cambodia.

## Author Contributions

R.E., J.P. and S.A. were involved in the conception and design of the study. R.E., A. Rai and A. Rumaseb performed the lab experiments. R.E., K.S.H, E.S., and H.T. performed data analysis. S.T., S.H., C.C., T.B., R.N.P., J.P. and S.A. provided critical clinical and metadata information and laboratory resources, and local epidemiological insight that were central to the study. R.E., J.P. and S.A. wrote the original draft of the manuscript. All authors reviewed and contributed to the final manuscript.

## Data availability

The genetic diversity of *Plasmodium vivax* using microhaplotype panel of sequence data is available in European Nucleotide Archive (ENA) and can be accessed under the BioProject PRJEB105391.

## Acknowledgements

We thank the patients who contributed their samples to the study, and the health workers and field teams who assisted with the sample collections.

## List of Supplementary Data Files

**Supplementary Data 1. Patient demographics and sample sequencing outcomes**

**Supplementary Data 2. Marker diversity and effective cardinality data**

**Supplementary Data 3. Within-host and population diversity data**

**Supplementary Data 4. Identity-by-descent data**

