## Supplementary information for "Temporal trends in *Plasmodium vivax* diversity in eastern Cambodia evidence declining transmission"

| Period (years) | Province | Sex | | Age (median, IQR) | Treatment | Number of samples |
|---|---|---|---|---|---|---|
| | | Male | Female | | | |
| **2014** | Rattanakiri | 65% (24/37) | 35% (13/37) | 20 (4-60) | DHA-PPQ | 37 |
| **2015** | Rattanakiri | 73% (33/45) | 27% (12/45) | 25 (8-60) | AS-MQ | 45 |
| **2019** | Mondulkiri | 96% (48/50) | 4% (2/50) | 27 (15-57) | AS-MQ | 50 |
| **2023** | Mondulkiri | 65% (32/49) | 35%(17/49) | 19 (9-54) | AS-MQ + PQ | 49 |

**Supplementary Table 1. Socio-demographic details from retrospective data of symptomatic *Plasmodium vivax* infections.** *Total number of *P. vivax* cases reported in the district. NA, not applicable. Case reports were not available in 2014. DHA: dihydroartemisinin, PPQ: piperaquine, AS: artesunate, MQ: mefloquine, PQ: primaquine

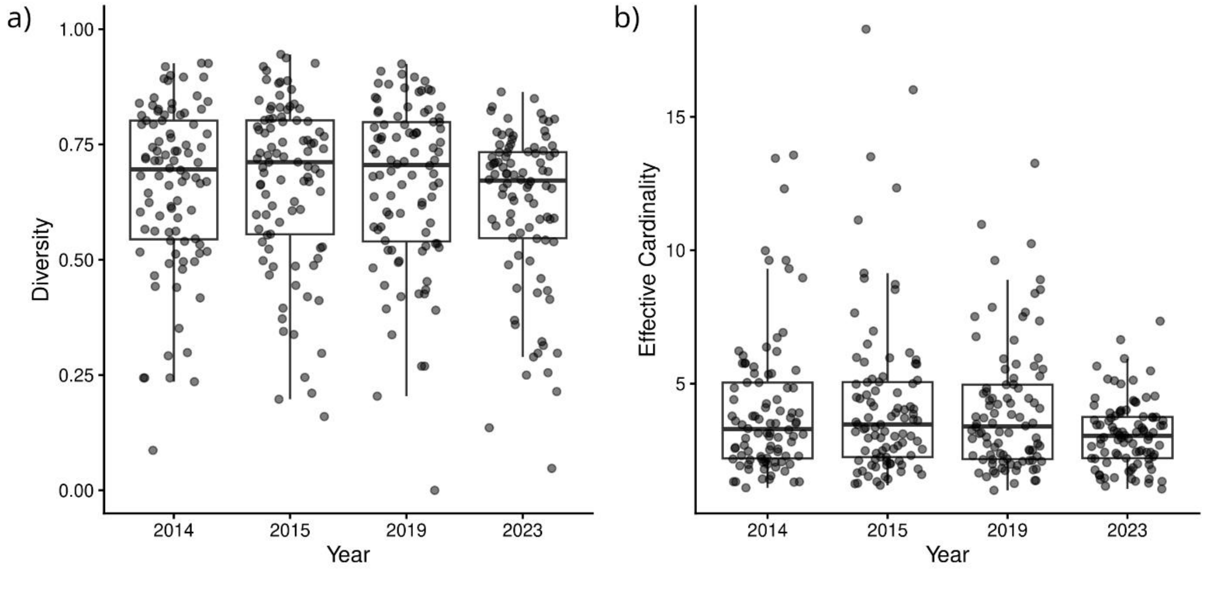


**Supplementary Figure 1. Marker-based allele diversity and effective cardinality in each of the study years.** Data is presented on the 92 high performance microhaplotype markers in the 110 monoclonal samples.

| Period (year) | % polyclonal cases | Mean eMOI* | Mean population HE** | Mean marker HE** | Mean marker Eff Card*** |
|---|---|---|---|---|---|
| 2014 | 41% (15/37) | 1.26 | 0.059 | 0.66 | 4.02 |
| 2015 | 22% (10/45) | 1.19 | 0.051 | 0.67 | 4.26 |
| 2019 | 48% (24/50) | 1.42 | 0.074 | 0.66 | 4.03 |
| 2023 | 4% (2/43) | 1.03 | 0.01 | 0.62 | 3.11 |

**Supplementary Table 2. Summary statistics on within-host, population-level and marker diversity.** *eMOI, effective multiplicity of infection. **HE, expected heterozygosity. ***Eff Card, effective cardinality.

a) b)

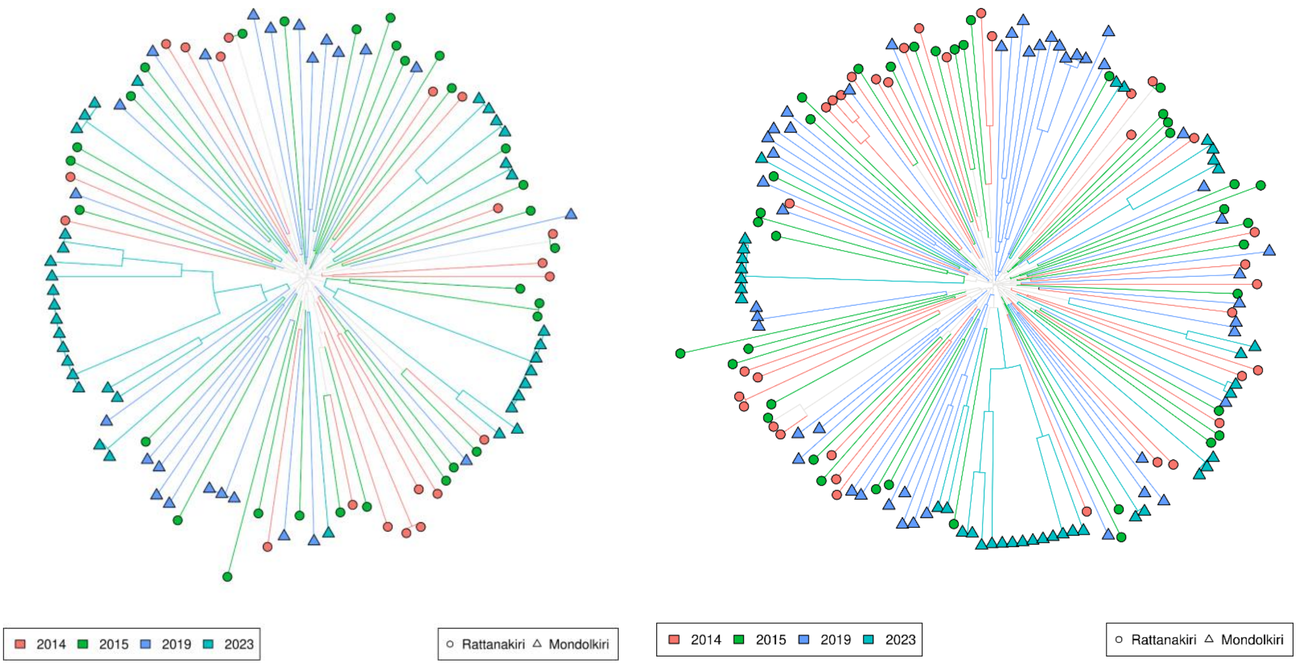


**Supplementary Figure 2. Intermixing of infections across the provinces and years of collection.** Panel a) presents a neighbor-joining tree generated on 110 monoclonal samples using 86 microhaplotype markers with no missing data in the sample set. Panel b) presents a neighbour-joining tree generated using data on 159 samples (monoclonal and polyclonal) with 86 microhaplotype markers. Major allele calls were used for heterozygote positions in the polyclonal infections.
